# COVID-19 Pandemic Response Simulation: Impact of Non-pharmaceutical Interventions on Ending Lockdowns

**DOI:** 10.1101/2020.04.28.20080838

**Authors:** Serin Lee, Zelda B. Zabinsky, Stephen Kofsky, Shan Liu

**Affiliations:** UW Healthcare Analytics Lab, Industrial & Systems Engineering Department, University of Washington, Seattle WA

## Abstract

As many federal and state governments are starting to ease restrictions on non-pharmaceutical interventions (NPIs) used to flatten the curve, we developed an agent-based simulation to model the incidence of COVID-19 in King County, WA under several scenarios. While NPIs were effective in flattening the curve, any relaxation of social distancing strategies yielded a second wave. Even if daily confirmed cases dropped to one digit, daily incidence can peak again to 874 cases without import cases. Therefore, policy makers should be very cautious in reopening society.

## I. Introduction

The novel coronavirus (COVID-19) pandemic is causing significant death tolls, economic losses and disruptions to the American society. As many parts of the U.S. are flattening the curve to control the current outbreaks and maintain hospital capacities through a combination of non-pharmaceutical interventions (NPIs) and expanded testing, the federal and many state governments are starting to coordinate efforts to ease restrictions and eventually end the lockdown. This project aims to explore the feasibility of gradually relaxing social-distancing orders when a vaccine is not yet available. Focusing on a large urban area (King County, WA), we simulated the outcomes of various opening-up scenarios for one year since the first infection started. This research will provide guidance for state and local governments in implementing policy changes regarding to lockdowns.

## II. Methods

### Model overview

We developed an agent-based simulation based on the open-source model FRED (a Framework for Reconstructing Epidemic Dynamics)^1^. Our current UWFRED model considered King County’s population distribution and daily contact pattern, COVID-19 transmission using SEIR model, current NPIs and was simulated for 365 days. We estimated new infection per day to measure the effects of easing social-distancing restrictions.

Baseline assumptions include:

- First infection in King County occurred on January 15^th^, 2020. Although the man lived in Snohomish County, he commuted to Seattle and thus might have impact on King County^2^. We also assumed this is the only non-local transmission. All other transmissions are community transmissions in King County.
- We divided patients into non-severe and severe groups. We assumed asymptomatic and mild symptomatic patients show similar patterns and considered them as non-severe cases. Severity of symptom depends on age^3^, and degree of severity does not influence infectivity.
- We assumed full immunity for people who recovered from COVID-19.

### Calibration procedure

We calibrated model parameters including COVID-19 transmissibility, daily contact patterns in household, neighborhood, school, and workplace, and probability of staying home when sick, targeting the basic reproduction number (R_0_) of COVID-19 which is known to be 2 to 3^4^. We used Latin hypercube sampling to form 1,000 candidate parameter sets. For each parameter set, we replicated the epidemic simulation 50 times to consider stochasticity of the simulation. After selecting the best calibrated parameter sets based on a goodness-of-fit criterion, we clustered them using complete linkage and Euclidean distance method to group results, determining six representative parameter sets (see Appendix 1).

### Interventions

We modeled three most common NPIs in the U.S. See Table 1.

**Table 1.** Simulated NPIs in King County

| Policy | Target | Period | Changes in contact pattern (among complied) | Compliance rate |
| --- | --- | --- | --- | --- |
| Case isolation at home<sup>1</sup> | Infected individual | onset of symptom +1 day ~ until recovery | 50% reduction in household contact<br>No school, workplace, neighborhood contact | Non-severe: default calibrated probability for stay home when sick + 10%<br>Severe: 100% |
| School closure | Students | 3/12~ 8/31 | No school contact | 100% |
| Social distancing<sup>2</sup> | Entire population | Differs by scenario | Same school/household contact<br>No workplace contact<br>No neighborhood contact | 30% - Low<br>60% - Medium<br>90% - High |

1. When 'case isolation at home' is activated, we assumed that people would reduce household contact and 10% more people stay home in non-severe case compared to the calibrated probability for staying home in King County. We assumed this intervention is effective from 02/28/2020, at the time which the first positive result in King County was announced to the public. 2. Each person and workplace comply to social distancing orders based on their default compliance rate. When a government issues a social distancing order, we categorized it as low, medium, and high. We assumed a percentage of workplace will remain closed and the same amount of people will not have neighborhood contact. The percentage is 30% for low, 60% for medium, and 90% for high orders.

Other possible NPIs, such as contact tracing and voluntary home quarantine, were not considered as they were either not being practiced in large scale in King County, or current social distancing already reflects such interventions.

### Scenarios for ending lockdowns

We simulated combinations of eight different levels of social distancing strategies that could be practiced after May 4^th^, the current final date of Washington government’s “Stay Home, Stay Healthy” emergency order, and a ‘do nothing’ scenario. We set interventions before May 4^th^ based on Washington state orders (Appendix 4). As extending school closure to the next school year is practically difficult, we assumed schools will reopen in September. See Table 2 and Figure 1.

**Table 2.**
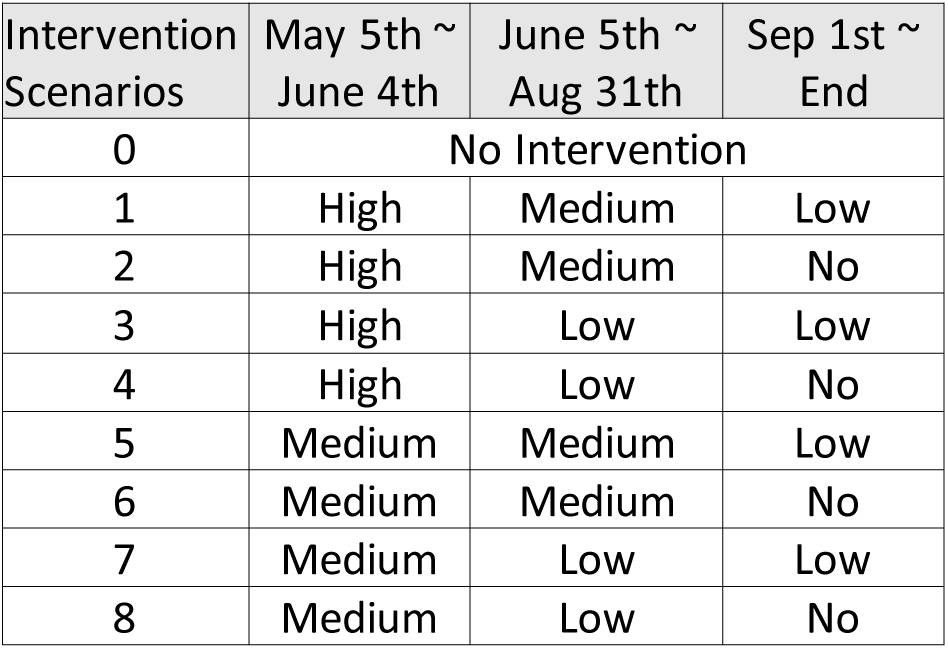
Scenarios

**Figure 1.**
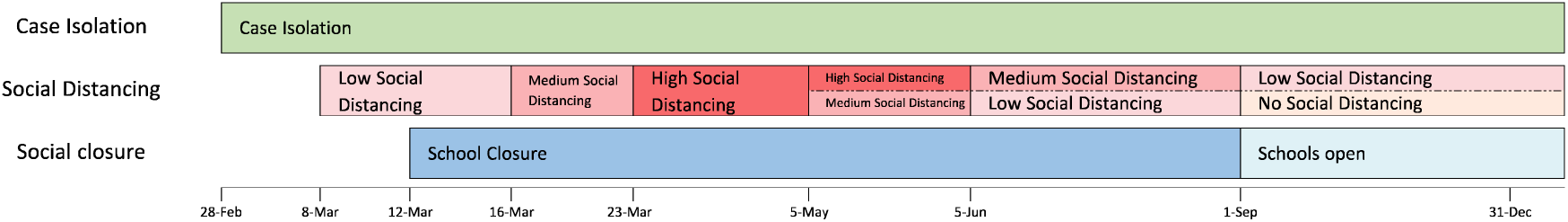
Diagram of Simulated Scenarios

## III. Results

### Calibration result

Out of 1,000 Latin hypercube parameter sets, 58 sets targeted R_0_ between 2 to 3. After clustering, six clusters had high similarity and low distance level (Appendix 1). Therefore, for each scenario, we used the centroid of the clusters and replicated 50 times for each centroid, totaling 300 simulations per scenario.

### Scenarios outcomes

Comparing Scenario 0 to Scenarios 1-8 shows that NPIs activated on mid-March were effective in flattening the curve and lowering total infection. As expected, Scenario 1, the most powerful and long-lasting social distancing, was most effective in reducing total infections from 41% to 15% of the population, compared to Scenario 0. New incidence peaked on March 18^th^ in Scenario 0 with 37,304 newly infected persons whereas other scenarios peaked on March 11^th^, with 18,881 persons. The ranking by effectiveness in reducing transmission is Scenario 1, 2, 5, 3, 6, 4, 7, 8, 0. See Figure 2.

**Figure 2.**
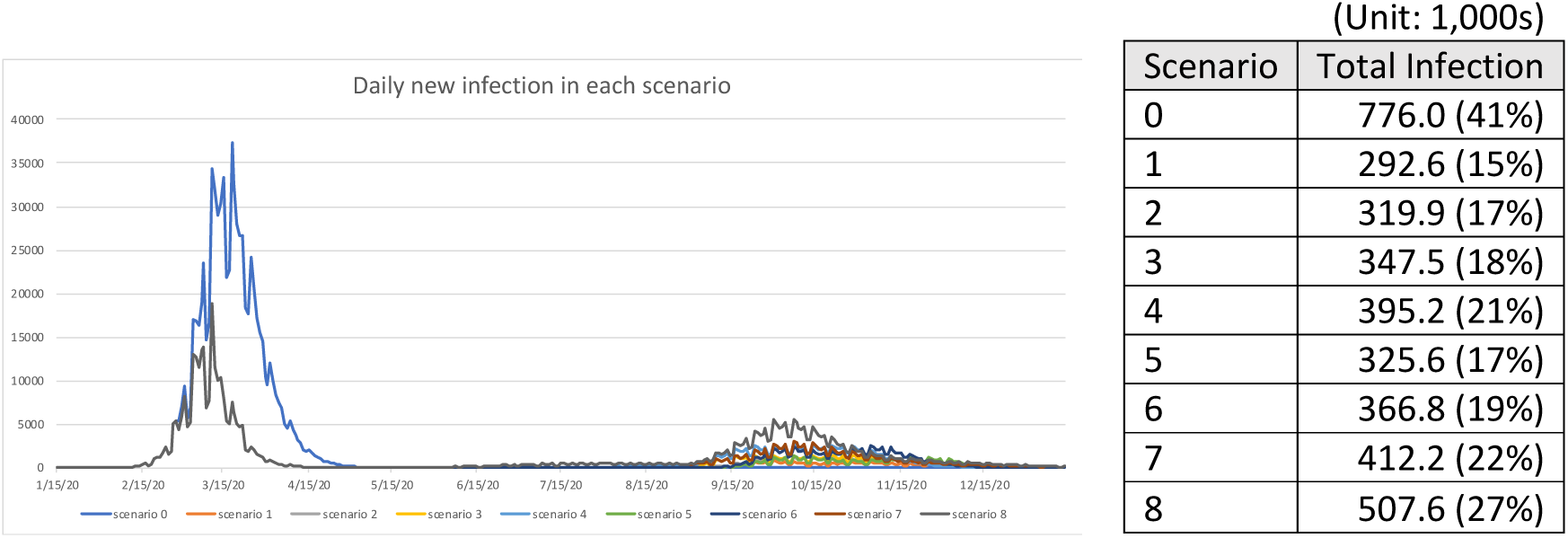
Simulated Scenarios Projections

However, Scenarios 1 to 8 all yielded a second wave. Even in Scenario 1, the best case, whose daily new infection dropped to 2 on June 4^th^ and remained around 20 per day until August, as soon as the social distancing strategies were relaxed and school opened, its incidence peaked again to 874 cases per day without import cases. It is noteworthy to look at the total infection since May 4^th^ for Scenarios 2 and 5. Although Scenario 2 totally reopened society from September 1^st^, the total infection is similar to Scenario 5 which retained low social distancing from September until the end of the simulation. This implies that extending one more month of strong social distancing until June 4^th^ could be as effective as retaining low social distancing in September. See Figure 3.

**Figure 3.**
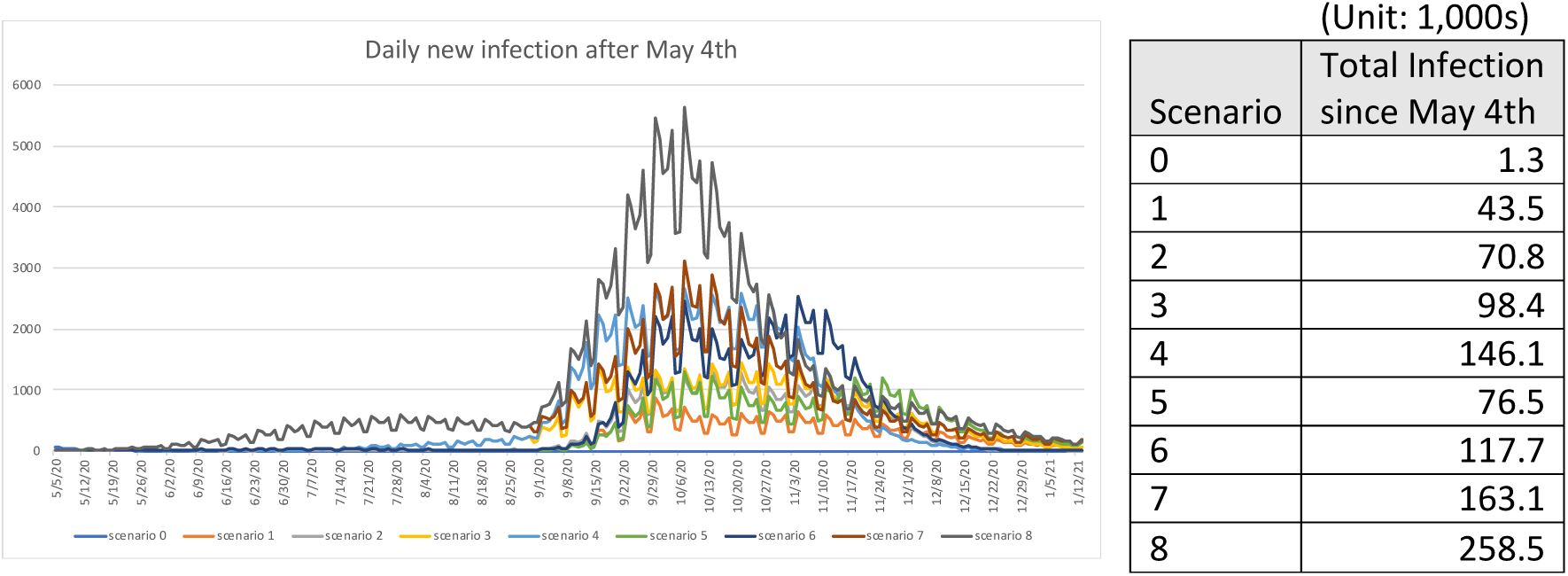
Projected Second Waves

In addition, the infections by place in Scenarios 1 and 2 (Figure 4) suggest that when social distancing is totally relaxed, peak workplace infection would double and neighborhood infection would more than triple compared to a low level of social distancing.

**Figure 4.**
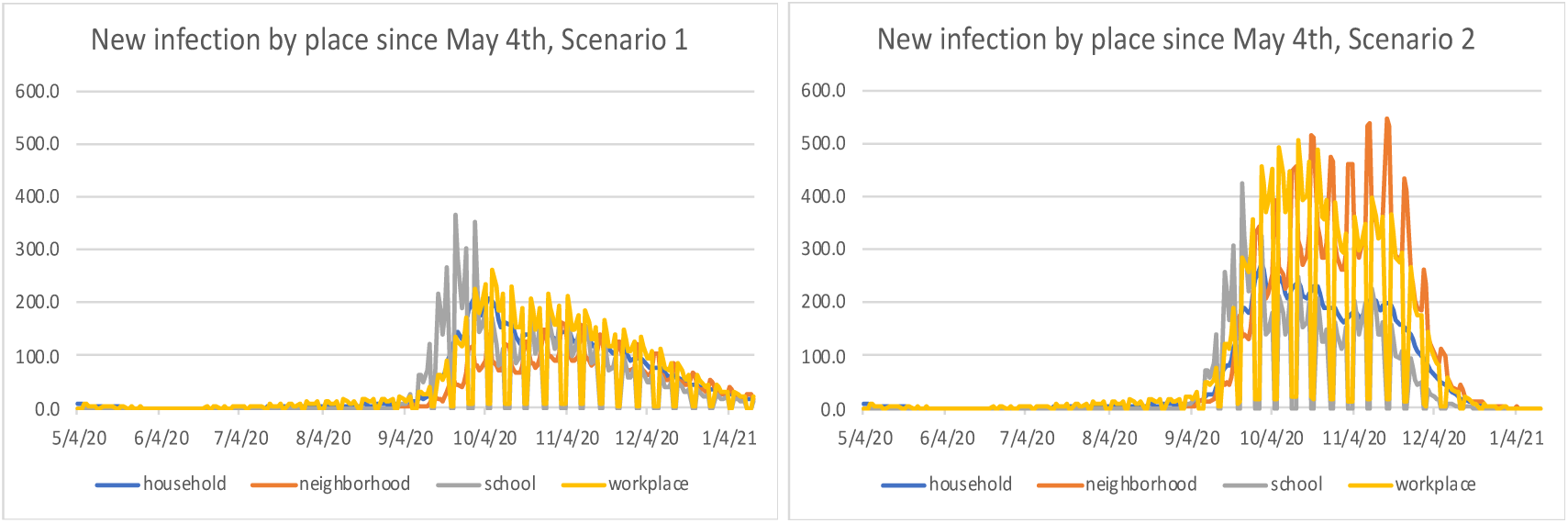
New Infections by Place

## IV. Conclusion

While reopening society is important in revitalizing economy, policy makers should be very cautious in reopening even if daily confirmed cases decrease to one digit or even zero due to low reporting rate.

The main limitation of this model is that in early transmission, daily incidences are much higher than reported cases. However, considering low reporting rate (12%) for symptomatic cases in the U.S^5^ especially until March, and COVID-19’s high prevalence in asymptomatic and mild cases^6^, the numbers might not be too much of an overestimate of the true incidence. In addition, we did not consider hospitalization and fatality as we focused on disease transmission. Considering 100% of severe patients self-isolate at home with only reduced contact in household, we assumed that the intervention performed similarly to hospitalization or death in terms of reducing disease transmission.

## Data Availability

All data used in the simulation model are from publicly available literature.

## Acknowledgement

This work has been supported in part by the National Science Foundation grant CMMI-1935403, and the University of Washington Graduate School Fund for Excellence and Innovation (GSFEI) Top Scholar Award.

## Appendix

### 1. Calibration result

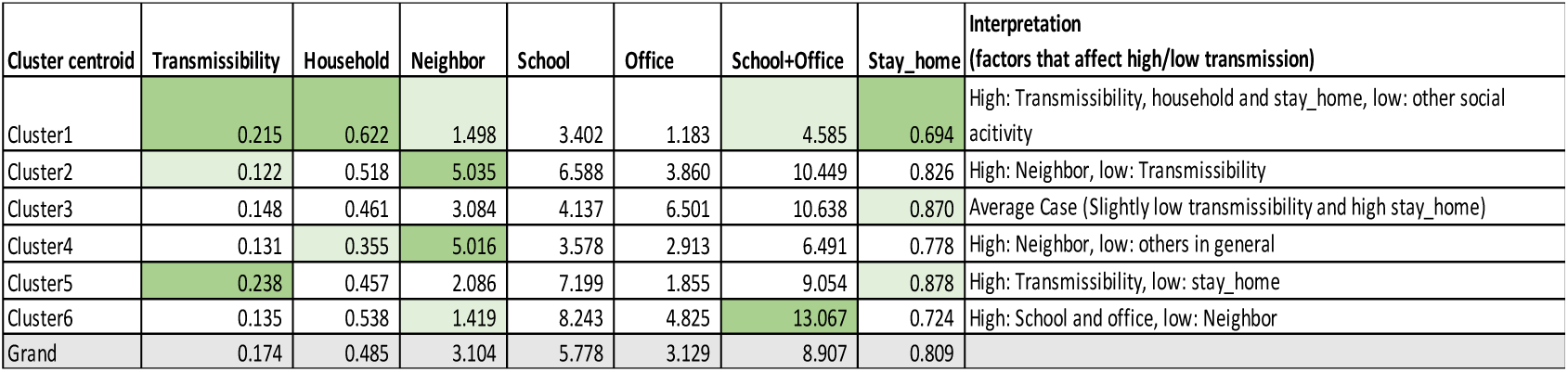

### 2. Simulation results by scenario and parameter cluster

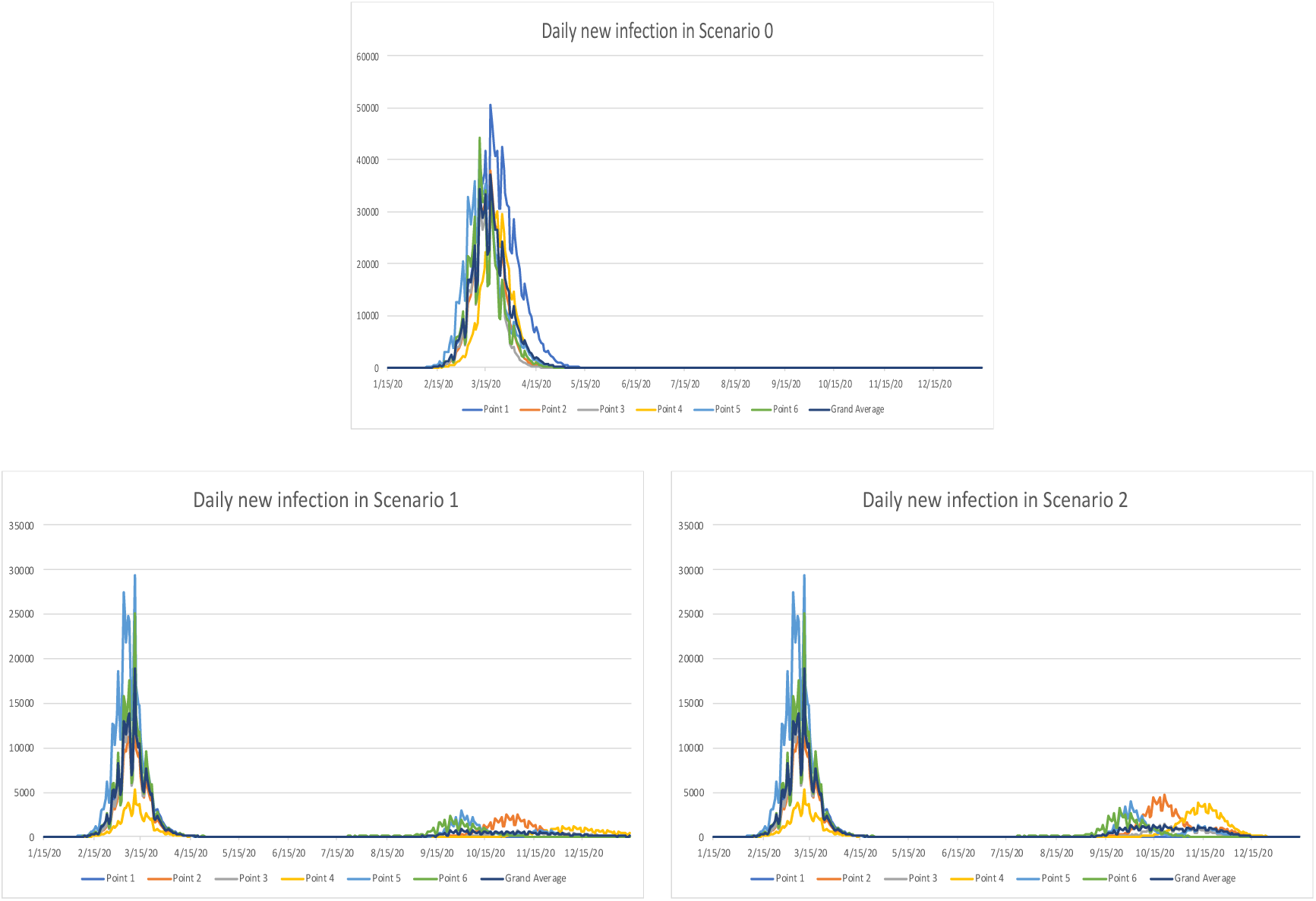

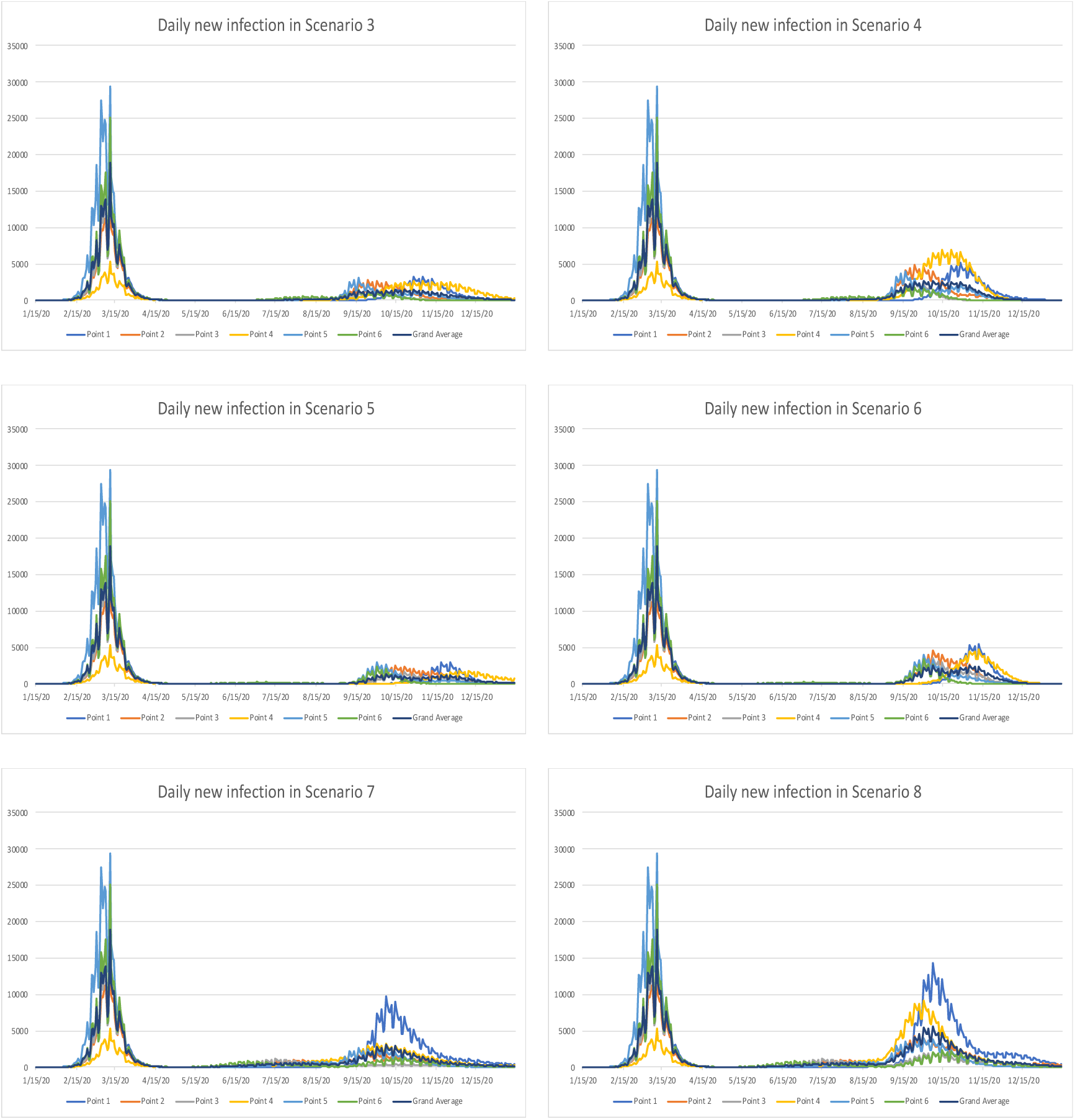

### 3. COVID-19 Simulation Input Parameters

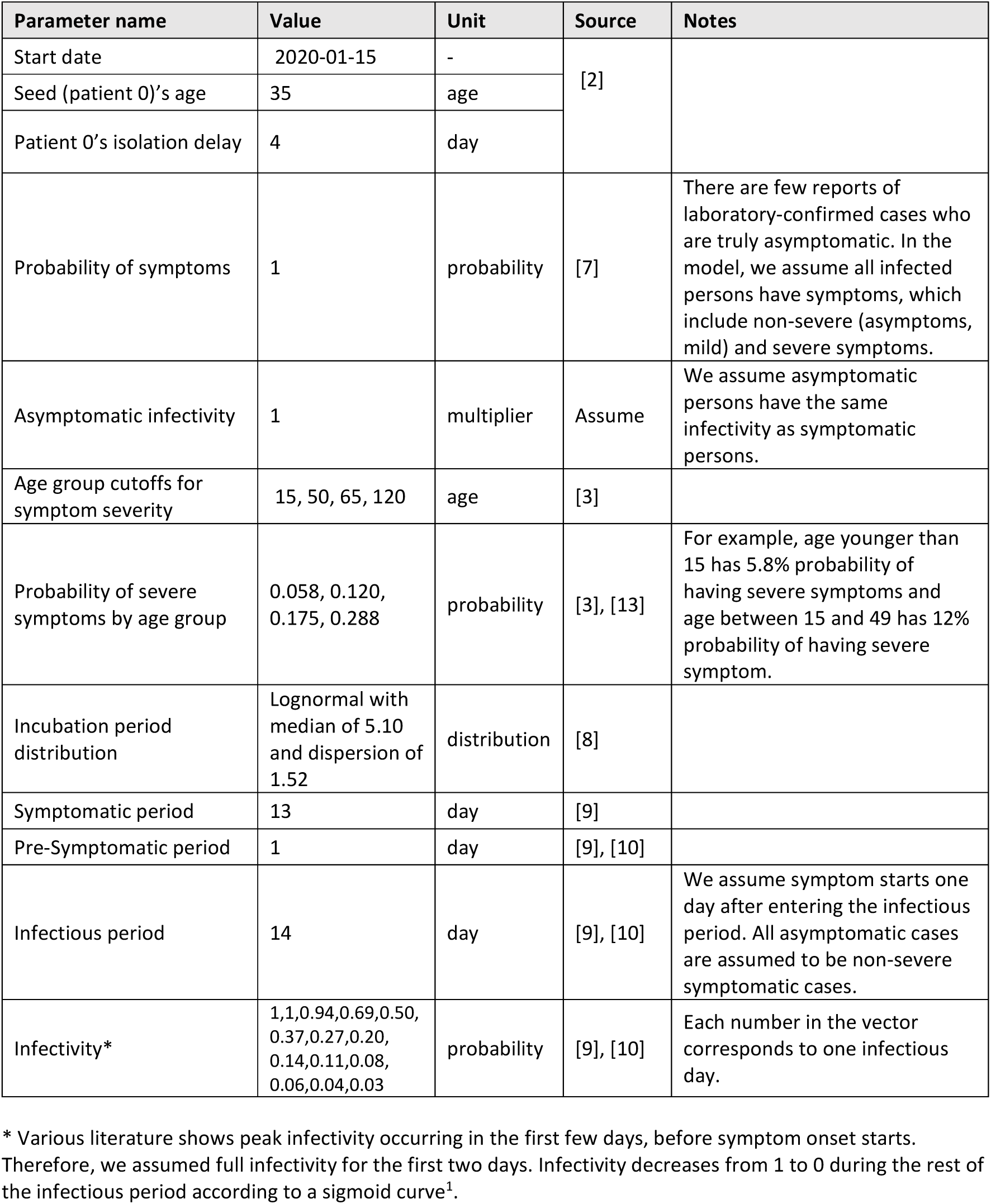

The SEIR structure is shown in the following figure^12^

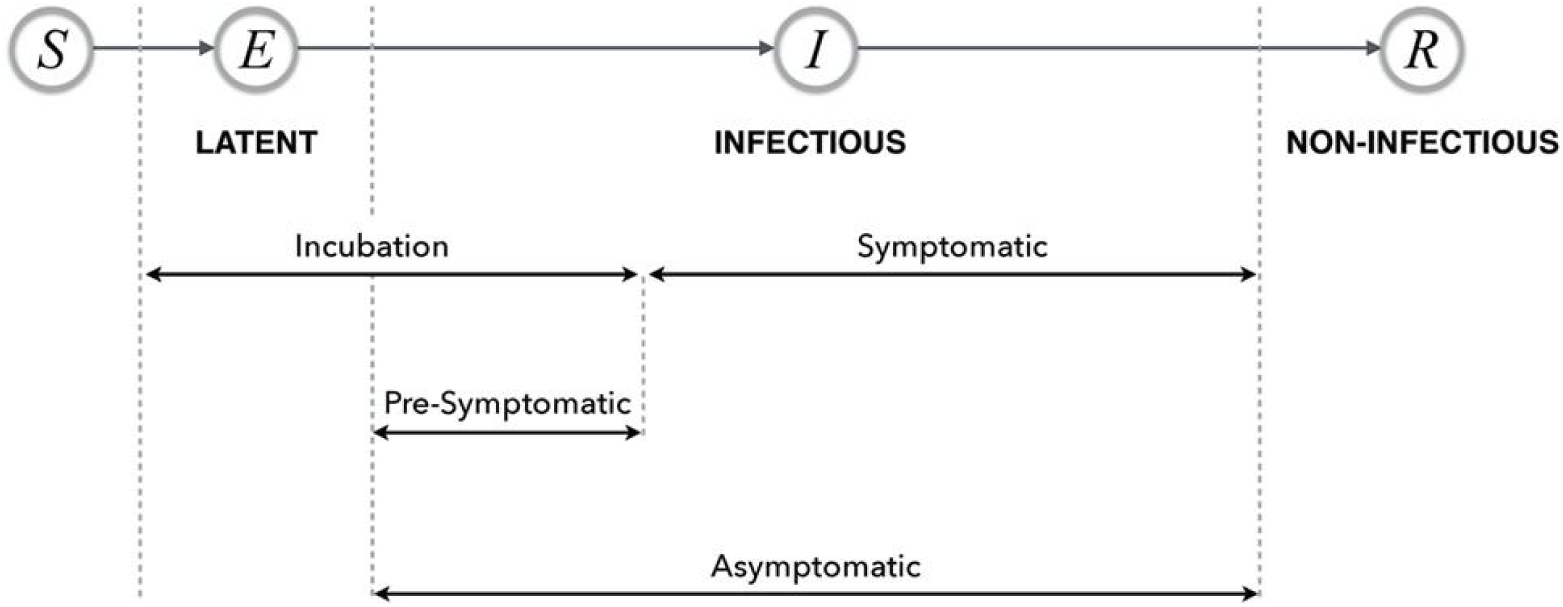

We simulated disease transmission following the procedure in FRED^1^.

a. In places other than household, for an infectious person i, the expected number of contact to transmit the disease to is:

> Contact_count = contacts_per_day(calibrated, by place) * transmissibility(calibrated)* infectivity(i)

For each contact j in Contact_count, the person j will get infected if fully susceptible. If the Contact_count is not integer, it is randomly rounded.

b If the place is household, pairwise transmission is simulated. For an infectious person i’s household member j, if the member is susceptible:

> Infection_prob = contact_prob(calibrated) * transmissibility(calibrated)* infectivity(i)* susceptibility(j)

If (Random number) < Infection_prob, the person j is infected.

### 4. Timeline of Washington state orders^11^

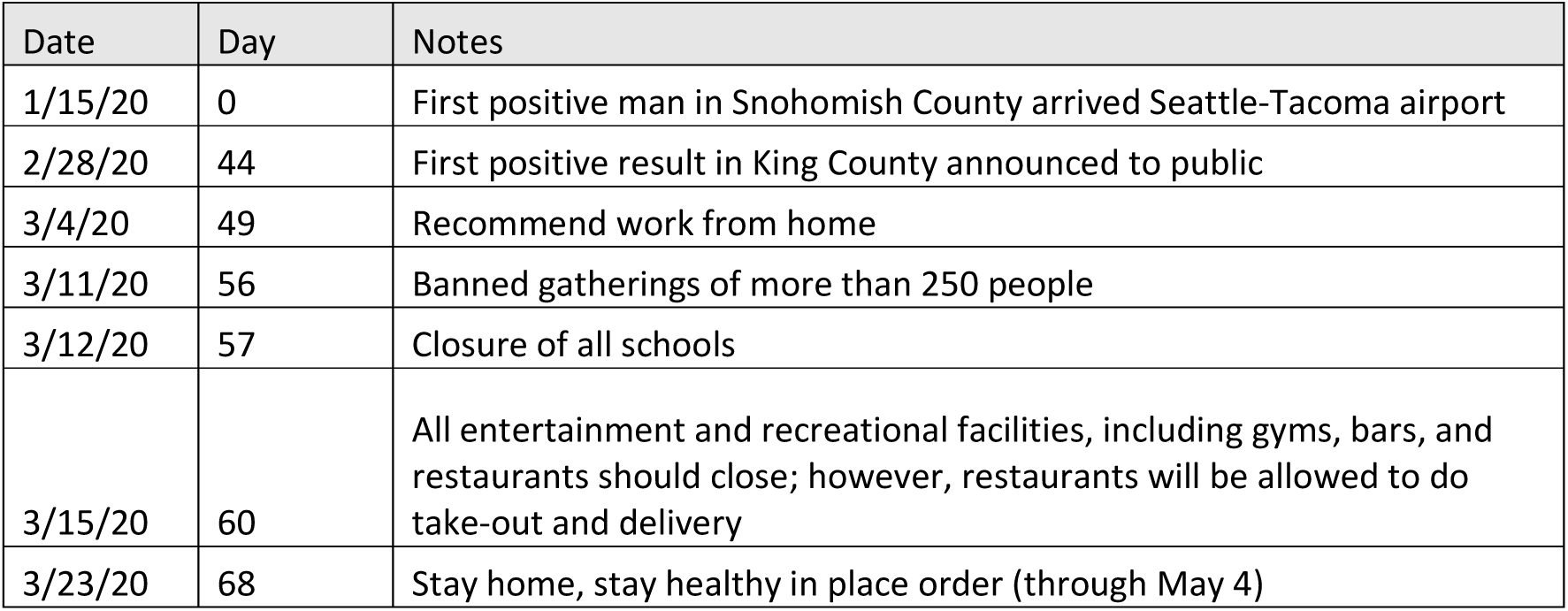

## References

1. Grefenstette JJ, Brown ST, Rosenfeld R, DePasse J, Stone NT, Cooley PC, Wheaton WD, Fyshe A, Galloway DD, Sriram A, Guclu H, Abraham T, Burke DS. 2013. FRED (a Framework for Reconstructing Epidemic Dynamics): an open-source software system for modeling infectious diseases and control strategies using census-based populations. BMC public health, 13: 940.

2. Holshue, M.L., DeBolt, C., Lindquist, S., Lofy, K.H., Wiesman, J., Bruce, H., Spitters, C., Ericson, K., Wilkerson, S., Tural, A. and Diaz, G., 2020. First case of 2019 novel coronavirus in the United States. New England Journal of Medicine.

3. Guan, W.J., Ni, Z.Y., Hu, Y., Liang, W.H., Ou, C.Q., He, J.X., Liu, L., Shan, H., Lei, C.L., Hui, D.S. and Du, B., 2020. Clinical characteristics of coronavirus disease 2019 in China. New England Journal of Medicine.

4. Liu, Y., Gayle, A.A., Wilder-Smith, A. and Rocklöv, J., 2020. The reproductive number of COVID-19 is higher compared to SARS coronavirus. Journal of travel medicine.

5. Russel, T., Hellewell, J. and Abbot, S., 2020. Using a delay-adjusted case fatality ratio to estimate under-reporting. Available at the Centre for Mathematical Modelling of Infectious Diseases Repository. https://fondazionecerm.it/wp-content/uploads/2020/03/Using-a-delay-adjusted-case-fatality-ratio-to-estimate-under-reporting-_-CMMID-Repository.pdf.

6. Gandhi, M., Yokoe, D.S. and Havlir, D.V., 2020. Asymptomatic Transmission, the Achilles’ Heel of Current Strategies to Control Covid-19. New England Journal of Medicine. https://doi.org/10.1056/NEJMe2009758

7. World Health Organization, 2020. Coronavirus disease 2019 (COVID-19): situation report, 73.

8. Lauer, S.A., Grantz, K.H., Bi, Q., Jones, F.K., Zheng, Q., Meredith, H.R., Azman, A.S., Reich, N.G. and Lessler, J., 2020. The incubation period of coronavirus disease 2019 (COVID-19) from publicly reported confirmed cases: estimation and application. Annals of internal medicine.

9. Young, B.E., Ong, S.W.X., Kalimuddin, S., Low, J.G., Tan, S.Y., Loh, J., Ng, O.T., Marimuthu, K., Ang, L.W., Mak, T.M. and Lau, S.K., 2020. Epidemiologic features and clinical course of patients infected with SARS-CoV-2 in Singapore. JAMA.

10. Anderson, R.M., Heesterbeek, H., Klinkenberg, D. and Hollingsworth, T.D., 2020. How will country-based mitigation measures influence the course of the COVID-19 epidemic? The Lancet, 395(10228), pp.931–934

11. Burstein, R., Hu, H., Thakkar, N., Schroeder, A., Famulare, M. and Klein, D., 2020. Understanding the impact of COVID-19 policy change in the greater Seattle area using mobility data. Institute for Disease Modeling. https://covid.idmod.org/data/Understanding_impact_of_COVID_policy_change_Seattle.pdf.

12. Milwid, R., Steriu, A., Arino, J., Heffernan, J., Hyder, A., Schanzer, D., Gardner, E., Haworth-Brockman, M., Isfeld-Kiely, H., Langley, J.M. and Moghadas, S.M., 2016. Toward standardizing a lexicon of infectious disease modeling terms. Frontiers in public health, 4, p.213.

13. Dong, Y., Mo, X., Hu, Y., Qi, X., Jiang, F., Jiang, Z. and Tong, S., 2020. Epidemiological characteristics of 2143 pediatric patients with 2019 coronavirus disease in China. Pediatrics.

